# Evolving concerns about the COVID-19 pandemic: A content analysis of free-text reports from the UK COVID-19 Public Experiences (COPE) study cohort over a two-year period

**DOI:** 10.64898/2026.04.16.26351013

**Authors:** Rhiannon Phillips, Fiona Wood, Anna Torrens-Burton, Clare Glennan, Paul Sellars, Sherina Lowe, Aleysha Caffoor, Britt Hallingberg, David Gillespie, Victoria Shepherd, Wouter Poortinga, Karin Wahl-Jorgensen, Denitza Williams

## Abstract

**Objectives:** Concerns about COVID-19 were a key driver of infection-prevention behaviour during the pandemic. The aim of this study was to gain an in-depth longitudinal understanding of the type and frequency of concerns experienced throughout the first two years of the COVID-19 pandemic.

**Design:** Content analysis of qualitative descriptions provided in a prospective longitudinal online survey as part of the COVID-19 UK Public Experiences (COPE) Study.

**Method:** At baseline (March/April 2020), when the UK entered its first national lockdown, 11,113 adults completed the COPE survey. Follow-up surveys were conducted at 3, 12, 18 and 24 months. Participants were recruited via the HealthWise Wales research registry and social media. Baseline surveys collected demographic and health data, and all waves included an open-ended question about COVID-19 concerns. Content analysis was used to identify the type and frequency of concerns at each time point.

**Results:** A total of 41,564 open-text responses were coded into six categories: personal harm (n=16,353), harm to others (n=11,464), social/economic impact (n=6,433), preventing transmission (n=4,843), government/media (n=1,048), and general concerns (n=1,423). The proportion of respondents reporting any concern declined from 75.3% at baseline to 65.8% at 24 months. Over time, concerns about personal harm increased (baseline 41.8% vs. 24-months 52.7%) whereas concerns about harm to others decreased (baseline 48.5% vs. 24-months 28.6%). Concerns about harm were also expressed in relation to clinical vulnerability, lack of trust in government/media, and perceived lack of adherence by others. These were balanced against concerns about wider social and economic impacts of restrictions.

**Conclusions:** Public concerns about COVID-19 evolved substantially over the first two years of the pandemic, reflecting changing perceptions of risk and responsibility. Monitoring concerns longitudinally is vital to help guide effective communication and behavioural interventions during future pandemics.

## Introduction

During pandemics perception of risk can strongly influenced people’s engagement in preventive behaviours such as mask-wearing, hand hygiene, social distancing, and vaccination uptake (1–14). Risk appraisal during a pandemic can range from feelings of mild concern to intense fear (15). Both internal and external factors such as easing and tightening of lockdown restrictions and introduction of widespread vaccination influence risk perception (5, 7, 16). Understanding how people’s concerns changed over time during the COVID-19 pandemic is vital for future pandemic planning to enable equitable and adaptive public health strategies to be developed and implemented (6).

‘Concerns’ are psychological appraisals of risk that form part of the affective aspect of risk perception, which incorporates emotional responses to a threat like worry and fear (17). Affective risk perception works alongside, but independently of, more reflective and analytical aspects of risk perception in determining behaviour (17). Quantitative longitudinal studies have indicated that overall perception of risk decreased over the time during the COVID-19 pandemic, corresponding with decreased engagement in protective action, but that affective risk perception was strongly associated with protective behaviour throughout (6, 18). A bi-directional relationship was observed between high perceived risk of COVID-19 and poorer mental health and well-being over the course of the pandemic (19). Qualitative studies also indicated that people’s concerns influenced infection-related behaviour and emotional well-being during the COVID-19 pandemic, and highlighted the importance of considering differences due to individual, demographic, contextual, cultural and geographic factors (20–24).

Monitoring evolving concerns can enable those planning pandemic-related communication to balance the need to convey seriousness with the need to prevent excessive fear or fatalism, which can erode resilience and social cohesion (25, 26). Longitudinal understanding of concerns can reveal emerging issues, such as worries about economic consequences or long-term health and well-being effects, that require policy adjustments beyond infection control alone (27–29). Large-scale studies of risk perceptions during the COVID-19 pandemic typically relied on broad quantitative measures of susceptibility of self/others to contracting infection, severity, risk of infecting others, and infection-related anxiety or fear (5–7, 15, 21, 30–33). However, these assessments do not fully illuminate the specific concerns that underlie the affective dimension of risk perception. Conversely, qualitative studies focused on providing in-depth insights into the views and experiences of specific populations and contexts during the pandemic, rather than providing broader insight into population-level shifts in the types of concerns reported in real time. As such, it is unclear how specific concerns about COVID-19 evolved over the course of the pandemic at a population level, which is essential in understanding how people balance concerns about harm from infection with concerns about the wider psychological, social and economic impact of the pandemic.

The aim of this study was to analyse open-ended qualitative responses from a large longitudinal UK community cohort to provide an in-depth understanding of how the type and frequency of concerns over the first two years of the COVID-19 pandemic.

## Method

### The COVID-19 Public Experiences (COPE) Study

Data included in this analysis were collected as part of the COVID-19 Public Experiences (COPE) project (5, 34, 35). COPE was a longitudinal mixed-methods study designed to examine public perceptions, experiences, and protective behaviour during the COVID-19 pandemic in the UK. The study comprised five online surveys conducted over a 24-month period between March 2020 and April 2022. Analysis of the quantitative survey data indicated motivational variables (fear of COVID-19, perceived susceptibility and perceived control) fluctuated over time as the disease and socio-political environment changed, decreasing overall by 24 months (6). Higher levels of fear, older age, lower perceived personal control over infection transmission, more trust in government and less trust in social media were associated with more infection–prevention behaviour (6). The current analysis extends previous work by providing a more in-depth understanding of specific concerns underpinning the observed quantitative changes in motivation over time. The COPE study received ethical approval from Cardiff Metropolitan University Applied Psychology ethics panel on 13.3.20 (Project reference Sta-2707).

### Theoretical approach

A social phenomenological approach was adopted in this study, using a data-driven process of understanding how people make sense of and interpret phenomena in their everyday world (36). We drew on established behaviour change theory to guide analysis and facilitate interpretation of data. Concerns, as part of affective risk perception is part of the ‘automatic’ component of motivation within the Capability, Opportunity and Motivation (COM-B) model of behaviour, which includes instinctive, drive-related and affective processes such as desires, fears and habits. The Plans, Responses, Impulses, Motives, Evaluations (PRIME) Theory of Motivation (16), elaborating on the motivational component of the COM-B, proposes that behaviour at any one moment arises from the strongest of potentially competing impulses and inhibitions (16). PRIME Theory postulates that affective motivation has a direct effect on behaviour but also influences behaviour via its impact on reflective motivation, which involves conscious decision-making, by creating a need or desire to plan and enact a particular behaviour (12). PRIME theory was relevant to developing theory-informed approaches to increase adherence to preventative measures during the COVID-19 pandemic (12). In the present study, we sought to use PRIME Theory and COM-B as a guide in developing insight into how different concerns, needs and impulses were balanced at different points in time.

### Study population and recruitment

A total of 11,113 adults living in the UK participated in the COPE study at the time of enrolment, which took place during the first national lockdown (13 March–13 April 2020)(34, 35). An early lineage of the wildtype SARSCoV2 carrying the D614G mutation was the dominant variant circulating in the UK at this time. Participants were recruited through social media advertisements (Facebook®, Twitter®, and Instagram®) and via the HealthWise Wales (HWW) research registry (37). The sample size was determined using a pragmatic approach, seeking to recruit sufficient numbers of people to enable us to understand a wide range of experiences in different geographical regions (urban/semi-rural/rural and varying levels of socio-economic deprivation) and demographic groups, allowing for attrition over time, while considering resources available and the need for a flexible and responsive approach to data collection in an unpredictable and rapildly changing context. At baseline, the majority of the 11,113 participants were female (69.2%), aged 51 years or older (68.3%), white British (95.8%), had a pre-existing medical condition (50.5%), and had received college (post-18) education (67.1%)(35). Further detail of demographic characteristics is provided in S1 Table.

Follow-up surveys took place at the following time points:

- 3-months, 20^th^ June 20 to 20^th^ July 2020, n=7,048, wildtype D614G variant dominant in UK, gradual easing of UK lockdown following the first wave of the pandemic
- 12-months, 12^th^ March 2021 to 13^th^ April 2021, n=5,535, Alpha variant dominant in UK, gradual easing of lockdown restrictions in UK after a second full UK-wide lockdown that had come into force in January 2021, a UK-wide COVID-19 mass vaccination program had commenced
- 18-months, 28^th^ September 2021 to 3^rd^ November 2021, n=4,242, Delta variant dominant in UK, most UK COVID-19 restrictions lifted and most UK adults had received at least one vaccination against COVID-19
- 24-months, 22^nd^ March 2022 to 28^th^ April 2022, n=3,827, Omicron variant dominant in UK, remaining COVID-19 restrictions being removed, UK government communications moved towards a ‘new normal’ where COVID-19 had become endemic

#### Materials and measures

The qualitative data analysed in this paper were participants’ responses to an open-ended question included in each of the COPE surveys. Free-text responses were collected at all five survey waves using the prompt:

> *“If you are worried about coronavirus (COVID-19), please tell us what your main concerns are.”*

These qualitative responses were analysed to explore how risk perceptions and related concerns evolved over time. The baseline survey also collected demographic and health-related data, including age, gender, highest educational qualification, ethnic group, presence of children under 18 years in the household, pre-existing medical conditions, and seasonal influenza vaccination within the preceding 12 months.

#### Analysis

Content analysis was performed on open-text responses relating to what people’s main concerns were (if they had any) at each time point (30). This approach enabled efficient analysis of large quantities of text into categories with similar meanings, allowed interpretation of meaning from the content and context of free-text using a naturalistic paradigm, and provided a quantitative indication of the prevalence of categories of concerns (30). An inductive analytic process was applied to the baseline data that involved familiarisation, generation of initial codes, combining codes into categories, reviewing categories, and determining significance of categories to generate a coding framework for the content analysis. The coding framework developed using the baseline data was applied deductively to the follow-up survey data to allow consistency in coding across time points. This was supplemented with further inductive analysis of concerns that did not fit into existing categories, producing new categories as required as the pandemic progressed.

Text responses were coded by a team of 11 researchers. Coders were an inter-disciplinary team with expertise in psychology, medical sociology, nursing, palliative care, biosciences, public health and health services research, were all females of working age, with a variety of social backgrounds and family situations. To enable effective team-based coding, a clear and supportive management structure was in place, the team met regularly to discuss analysis and interpretation, detailed guidance and reference materials (codebook, written guidance and training materials) were provided, and coders were supported in building up skills gradually (38). Comments were coded in Microsoft Excel to accommodate the volume of data and coding by multiple researchers. Comments could be coded into multiple categories if required.

A random sample of 10% of the baseline data were verified by a second member of the research team (RP) to check the consistency and accuracy of coding across the team. Comments coded within the ‘general concerns’ category at all time points were independently coded by a second member of the team (RP/ATB) to assess whether they could be re-coded within one of the existing codes and to identify the need for new codes. Comments written in Welsh were coded directly from the text by a bilingual Welsh/English member of the coding team (RP). On completion of coding and quality assurance, the frequency of codes assigned to each main category and sub-category at each time point was counted (30).

## Results

In total, 41,568 open text comments were written by survey participants to describe their main concerns about COVID-19. Concerns were provided by 8,372 of 11,113 (75.3%) people at baseline, 5,330 of 7,048 (75.6%) at three-months, 3,823 of 5,535 (69.1%) at 12-months, 2,928 of 4,242 (69.0%) at 18-months, and 2,518 of 3,827 (65.8%) at 24-months. This overall reduction in the proportion of people reporting concerns was consistent with the overall decrease in fear of COVID-19 observed using quantitative data from the COPE cohort (6).

The six main categories of concerns identified through content analysis covered: 1. Personal harm (n=16,353 comments); 2. Harm to others (n=11,464 comments); 3. Social and economic impacts (n=6,443 comments); 4. Preventing the spread of COVID-19 (n=4,838 comments); 5. Government and media (n=1,048 comments), and 6. General concerns (n=1,423 comments). Fig 1 illustrates the change in percentage of participants reporting one or more concerns in each main category at each survey.

**Fig 1.**
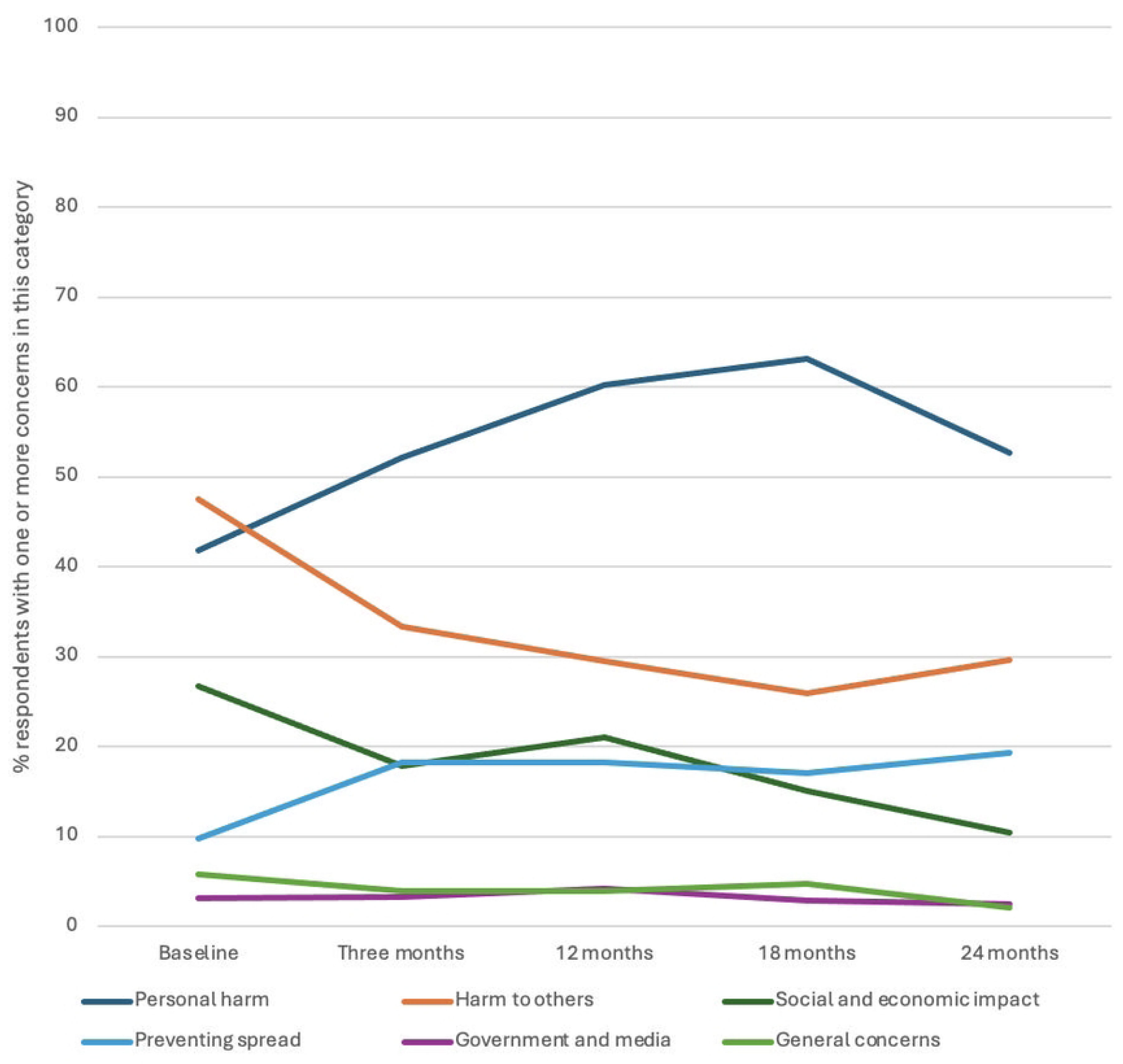
Percentage of respondents reporting main categories of concerns at each survey time point

Each main category is further divided into sub-categories, which capture the breadth and nuance of participants’ concerns across five survey time points, illustrating how priorities and fears shifted over the two-year study period. A total of 30 sub-categories were identified (Table 1). Individuals who provided a description of their concerns in each survey reported between one and 10 sub-categories of concern, with the median being two sub-categories of concerns reported by individuals at baseline, 12-months and 18-months, and a median of one sub-category of concerns at 3-months and 24-months. A table of exemplar quotes in main and sub-categories at each survey time point is provided in S2 Table.

**Table 1:**
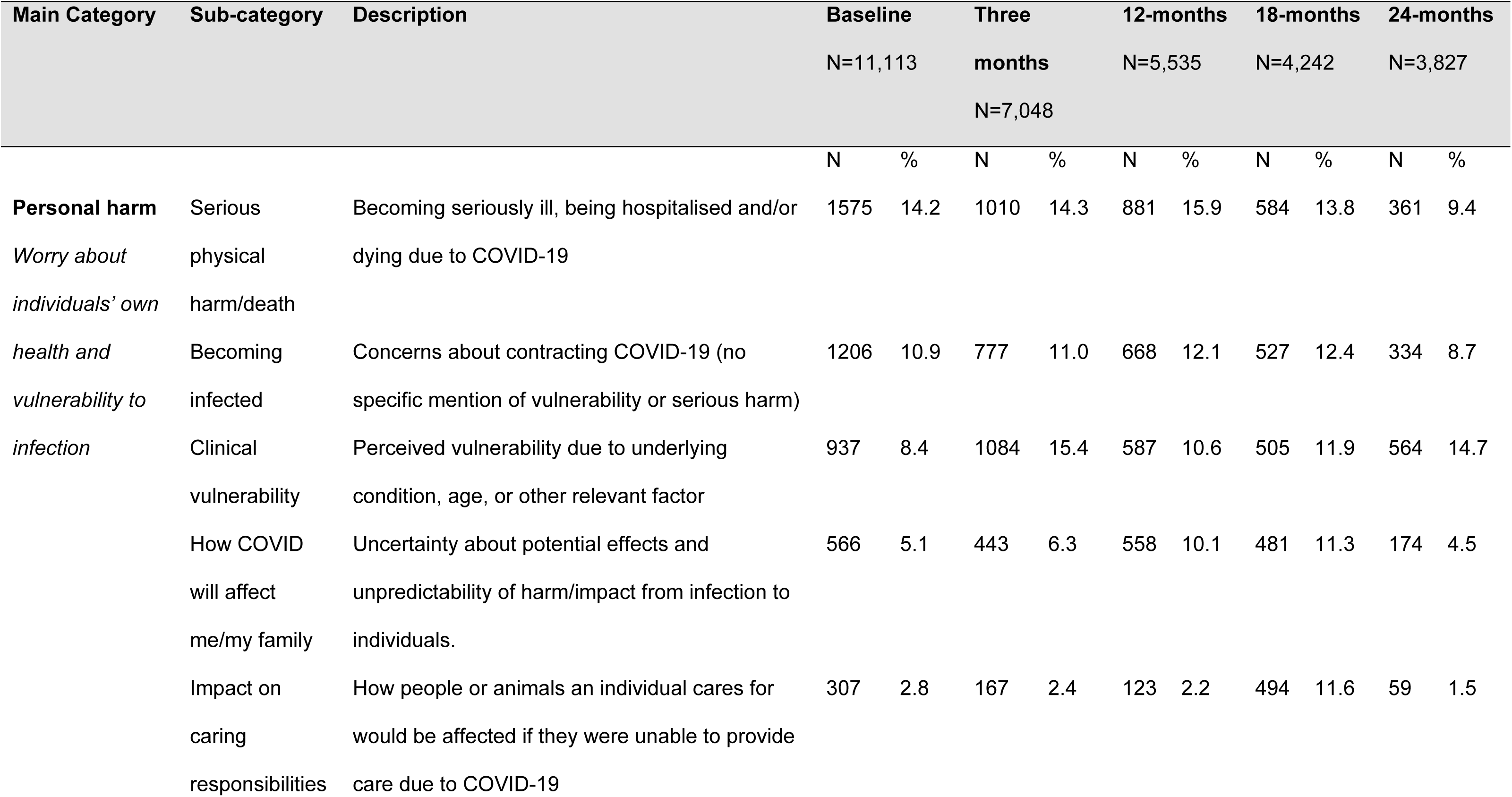
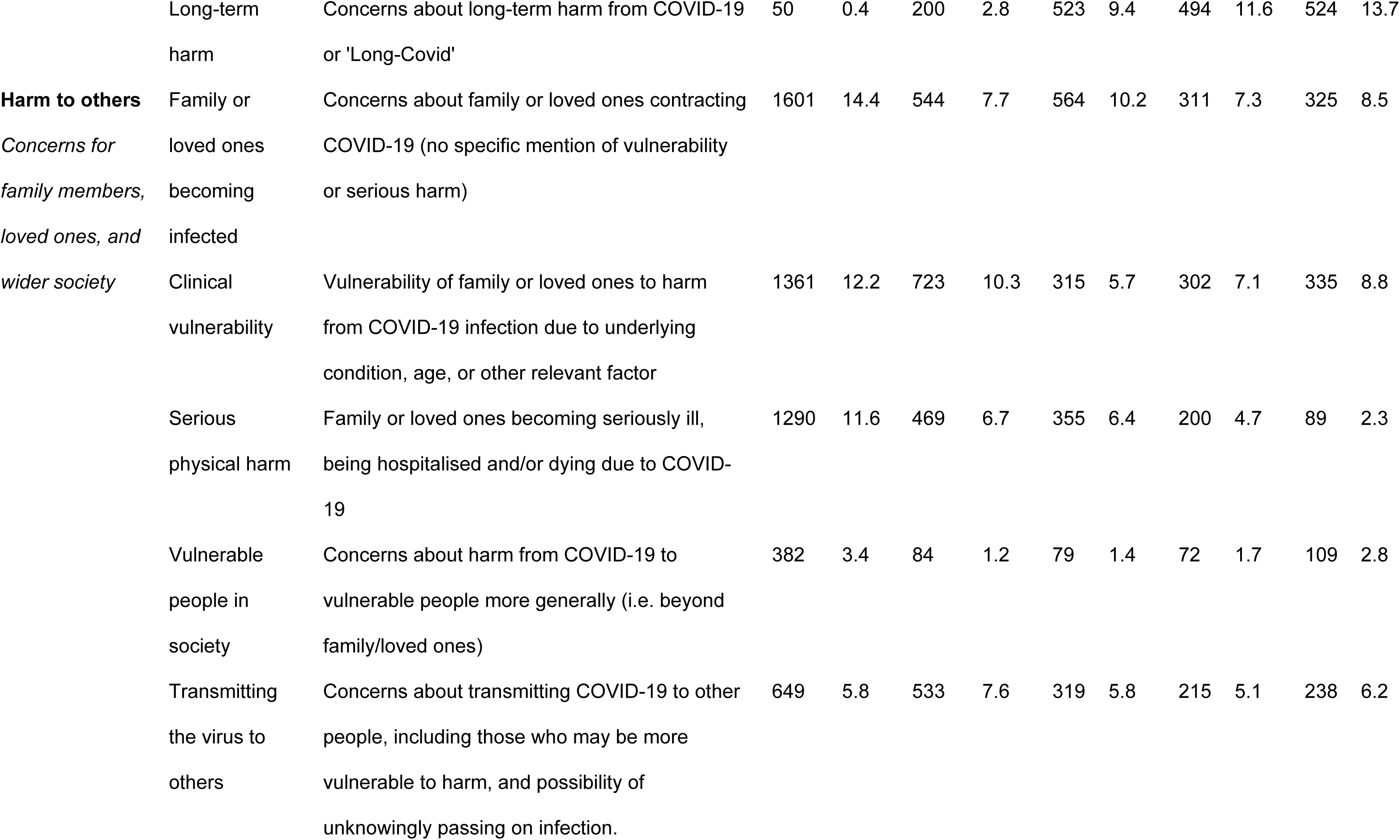
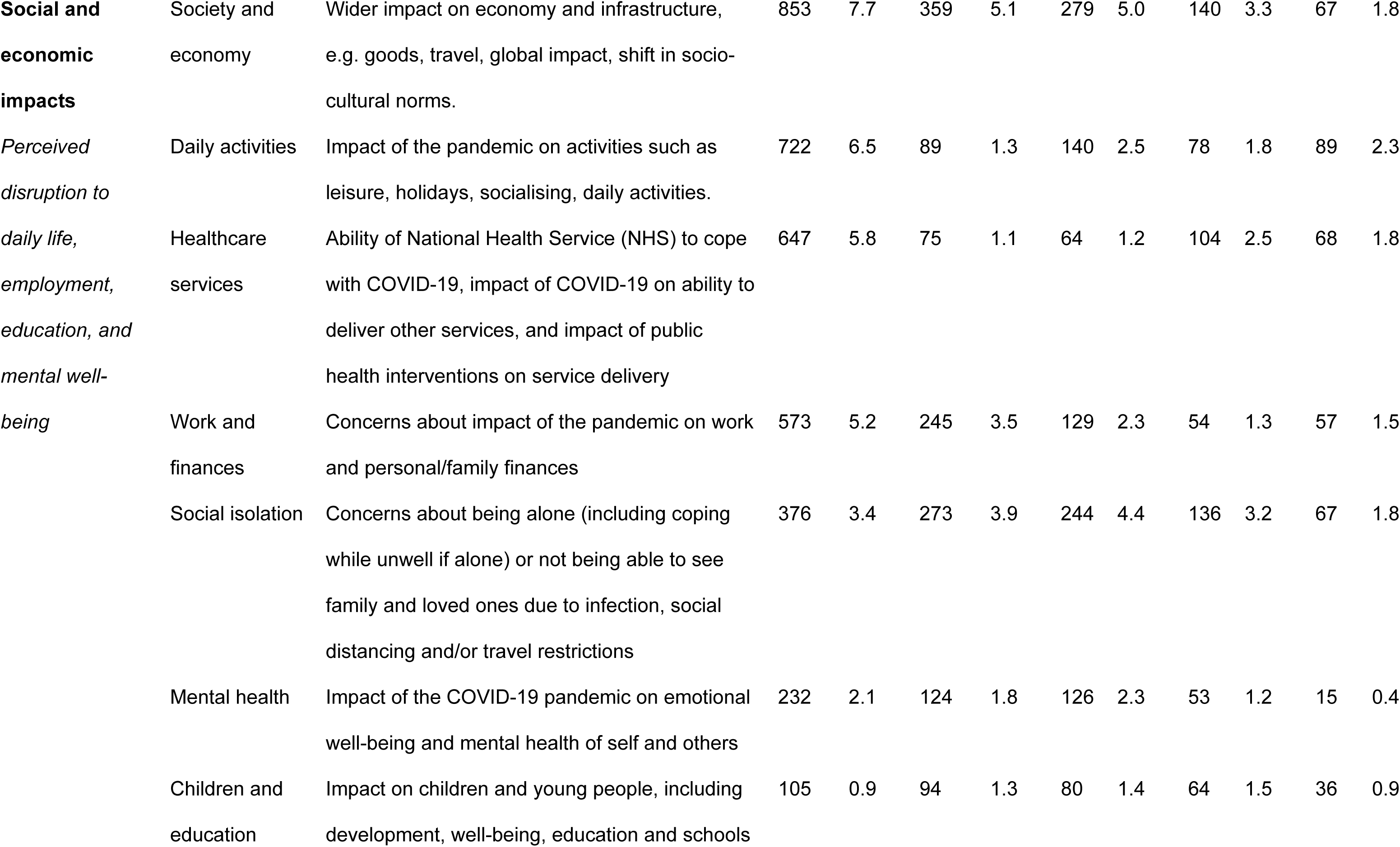
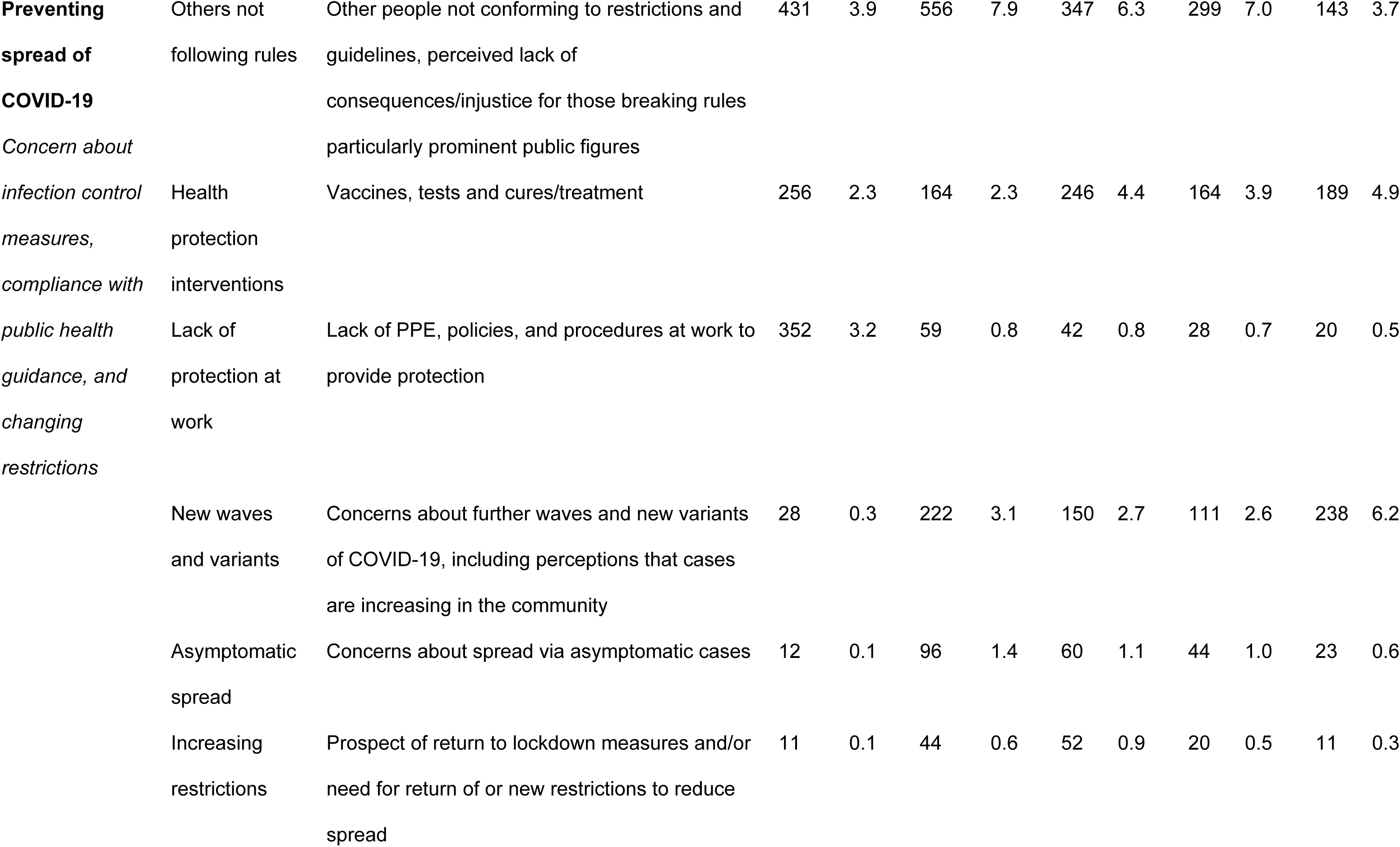
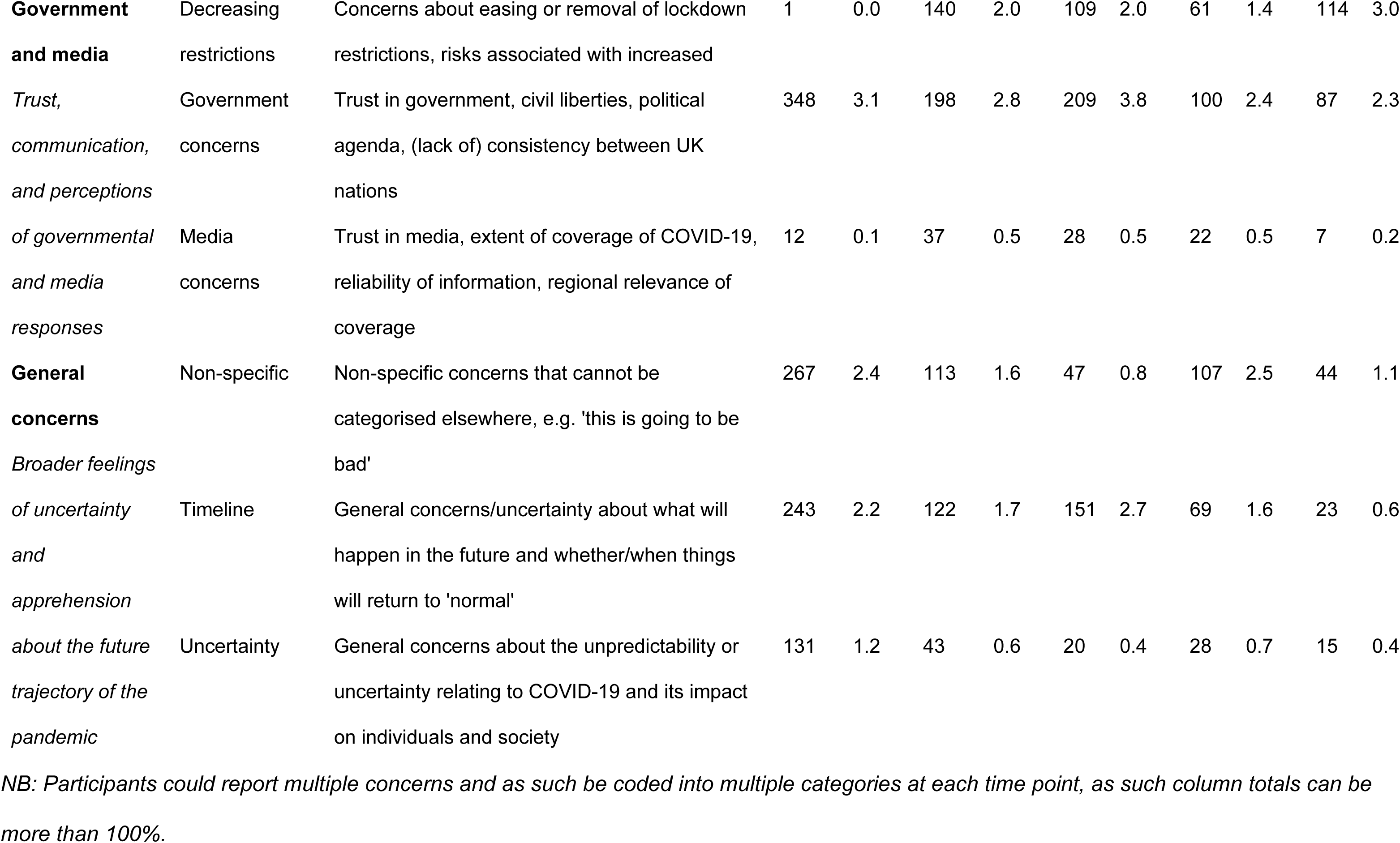
Number and % reporting specific sub-categories of concerns at each survey time point.

### Personal harm from COVID-19

Personal harm from COVID-19 was the most common concern at all time points, with one or more concerns within this category reported by 4,641/11,113 (41.8%) respondents at baseline, 3,681/7,048 (52.2%) at 3-months, 3,340/5,535 (60.3%) at 12-months, 2,675/4,242 (63.1%) at 18-months, and 2,016/3,827 (52.7%) at 24-months. Early in the pandemic, concerns centred on severe illness, hospitalisation, breathlessness, and death were prominent.

> *“I’m worried about being in pain and maybe being in hospital alone. I’m worried about dying and leaving my family.”*
>
> (P1, female, age 41-50, pre-existing condition(s), baseline)

Despite ongoing concern about personal harm from COVID-19, fears about serious harm or death decreased over time (14.2% at baseline vs. 9.4% at 24 months).

Concerns about clinical vulnerability peaked at three and 24 months, when population-level government protections were being reduced/removed. Clinical vulnerability was a major driver of perceived personal risk, particularly among those with existing health conditions.

> *“I’ve been terrified that Covid could kill me (I’ve been in ICU previously with my asthma)”*
>
> (P2, female, age 31-40, pre-existing condition(s), 12-months)

Participants worried about contracting COVID-19 even without severe consequences, due to the unpleasant experience of illness and disruption to daily life.

> *“Although I did not require hospital treatment, COVID-19 completely floored me. I was terribly poorly for a couple of weeks and then continued to suffer with the longest lasting ‘flu’ I’ve ever experienced.”*
>
> (P4, female, age 51-60, no pre-existing condition, 24-months)

The perceived unpredictability of the illness heightened concern about personal and familial outcomes.

> *“No two people are affected in the same way and no way of knowing how the infection would develop in myself & family.”*
>
> (P5, female, age 71-80, pre-existing condition(s), 18 months)

Over time, worries about long-term health consequences, particularly long-covid, became more prominent as people balanced reduction in restrictions with ongoing risks of infection.

> *“The number of cases going up and people starting to mix I am concerned that either I, or more vulnerable friends and family, will get Covid and be very ill or get long-covid. We can’t stay cooped up forever, but it is still a dangerous disease even though we are vaccinated”*
>
> (P6, female, age 41-50, pre-existing condition(s), 18-months)

Across all stages of the pandemic, participants worried about how illness might impair their ability to care for dependants, including children and vulnerable adults.

> *“I am also concerned that my wife and I will catch it, and suffer the symptoms of it, at the same time and be unable to effectively look after our children.”*
>
> (P7, male, age 31-40, pre-existing condition(s), baseline)

> *“I am an unpaid carer for our adult autistic son. If we become unwell, there is no back up plan. If he becomes unwell, he cannot tolerate pain or hospitalisation. We have no support.”*
>
> (P8, female, age 61-70, pre-existing condition(s), 18-months)

Concerns about harm from COVID-19 persisted for some even after vaccination, indicating that immunisation reduced but did not eliminate perceived threat.

> *“I have severe asthma and bronchiectasis. Although I am double vaccinated and getting my booster on November 5^th^, I am still too nervous to go outside unless I really have to.”*
>
> (P3, female, age 61-70, pre-existing condition(s), 18 months)

> *“The fact that it [COVID-19] has not gone away and even though I have been fully vaccinated I know I could catch it and be fairly ill.”*
>
> (P34, female, age 71-80, pre-existing condition(s), 24 months)

### Harm to others from COVID-19

Concerns about harm to others declined over time, with 5,283/11,113 (47.5%) reporting one or more concern in this category at baseline, 2,353/7,048 (33.4%) at 3-months, 1,632/5,535 (29.5%) at 12-months, 1,100/4,242 (25.9%) at 18-months, and 1,096/3,827 (28.6%) at 24-months. Early worries focused on family members or loved ones becoming seriously ill or dying, particularly those perceived as clinically vulnerable due to age or pre-existing conditions.

> *“My main concern is not for me, I am fairly fit and healthy and had a dose of a fluey/coldy bug before Christmas, I am very afraid for my partner considering that the last bout of flu that he had, he ended up in hospital for three weeks due to pneumonia and a heart attack induced by the pneumonia.”*
>
> (P10, female, age 41-50, no pre-existing conditions, baseline)

> *“A terrible death for myself and my family members.”*
>
> (P11, male, age 61-70, pre-existing conditions, 12 months)

Participants also worried about transmitting the virus to vulnerable relatives or partners.

> *“My concern is catching it and passing it on to my partner as he is in a high-risk category. I think I would be ok but he could genuinely die.”*
>
> (P13, female, age 41-50, no pre-existing conditions, baseline)

These concerns were intensified by perceptions that others were behaving irresponsibly and by the removal of public health protections.

> *“I remain worried that I or someone I love will experience serious health problems as a result of catching the virus. I believe that with the end of free testing and the push to return to normality I have very little control now over exposure to infection.”*
>
> (P12, male, age 41-50, no pre-existing condition, 24-months)

Beyond immediate family, concerns for vulnerable groups within communities and globally were commonly reported.

> *“Worried about the weakest members of my community, worried about local business closing down, worried about permanent damage the virus might cause”*
>
> (P14, female, age 31-40, pre-existing condition, three-months)

### Social and economic impacts

Participants reported a wide range of concerns about the broader societal consequences of the pandemic, with 2,971/11,113 (26.7%) respondents at baseline reporting a concern in this category, 1,259/7,048 (17.9%) at 3-months, 1,164/5,535 (21.0%) at 12-months, 640/4,242 (15.1%) at 18-months, and 399/3,827 (10.4%) at 24-months, indicating that these concerns were more common during periods when restrictions were more stringent. At baseline, worries focused on infrastructure and access to essential supplies, especially during periods of panic buying and potential self-isolation.

> *“I worry not about the illness but about the way people are shutting themselves away and panic buying food. It will be a disaster for an already fragile economy.”*
>
> (P15, female, age 51-60, pre-existing condition(s), baseline)

Concerns extended to the wider social, economic, and cultural impacts of COVID-19 and associated public health interventions, including effects on health workers, the economy, and vulnerable populations.

> *“I’m mainly worried about the effects on society, health workers, the economy and the most vulnerable people”*
>
> (P16, male, age 41-50, no pre-existing condition, 12 months)

> *“I am very concerned at the strain the pandemic has put on health and social care workers and the way the education and social development of children and young people have been affected.”*
>
> (P17, female, age 61-70, no pre-existing condition, 24 months)

Disruption to daily life and cancelled plans were frequently mentioned, often linked to declining mental well-being.

> *“My concerns focus on the disruption to my life and the lives of the people I love. Plans for the year have been cancelled and this can have an effect on my mental health.”*
>
> (P18, female, age 31-40, no pre-existing conditions, baseline)

Financial insecurity and employment instability were key concerns, ranging from temporary income loss to potential long-term business collapse.

> *“That I will lose my business, due to an economic collapse and/or collapse of the delivery infrastructure (I run an online shop)”*
>
> (P19, female, age 41-50, no pre-existing condition, baseline)

> *“How I would financially support myself if I had to self-isolate and could not work or if I were off work due to sickness with it.”*
>
> (P20, female, age 31-40, pre-existing condition(s), 24-months)

Concerns about the NHS focused on its capacity to manage COVID-19 cases and the implications for access to routine healthcare.

> *“That the NHS won’t cope. That someone I care about will die, either because of COVID-19 or something else because hospital too full.”*
>
> (P21, female, age 51-50, no pre-existing conditions, baseline)

> *“I’m more worried about access to health care for existing chronic health conditions than I am about Covid.”*
>
> (P22, female, age 51-60, pre-existing condition, three-months)

Social isolation was a prominent concern, especially regarding separation from loved ones and the well-being of those living alone or at high risk.

> *“My biggest worries are for my family with my father and two grandparents considered high-risk for COVID19. I am worried both for their health and their mental well-being if they were to become isolated for an extended period of time.”*
>
> (P23, male, age 18-10, no pre-existing conditions, baseline)

Geographical distance compounded these concerns for some families.

> *“Our parents live 180 miles+ away in England and the Welsh Government limits us to 5 miles radius for travel. So, unless we do a ‘Dominic Cummings’, we are largely on our own, as all our other friends have children who have to social distance.”*
>
> (P24, male, age 31-40, pre-existing condition(s), three-months)

The reference to ‘Dominic Cummings’ related to a prominent government advisor who travelled 264 miles with his family despite having symptoms of COVID-19 in May 2020, which was widely reported in the UK mainstream media (28).

Some participants reported concerns about mental health impacts, including anxiety, loneliness, and exacerbation of pre-existing conditions.

> *“Concerned about pressure on NHS staff and Carers who might be suffering from PTSD. Concerned about mental well-being of friends who live alone.”*
>
> (P25, female, age 71-80, pre-existing condition(s), three months)

Concerns about children and young people during lockdown periods focused on education, development, and well-being.

> *“I am most concerned about the severe negative effects on my grandchildren’s lack of education and socialising, and on the welfare of my family than I am about becoming physically unwell.”*
>
> (P26, female, age 61-70, pre-existing condition(s) three months)

When schools reopened, worries shifted to infection risks in educational settings, particularly for clinically vulnerable families.

> *“I have 4 children who have to go to school - 2 in a very large secondary and 2 clinically extremely vulnerable children whom the school and council have said they will be fined for non-attendance even though they have letters to shield till 31st March. The fallout if any of us catching Covid will be very tough, but catastrophic if I as the main person who cares for the rest of the household becomes ill.”*
>
> (P27, female, age 41-50, pre-existing condition(s), 12 months)

### Preventing spread of COVID-19

Concerns in this category became more common as time progressed, reported by 1,091/11,113 (9.8%) respondents at baseline reporting a concern in this category, 1,281/7,048 (18.2%) at 3-months, 1,006/5,535 (18.2%) at 12-months, 727/4,242 (17.1%) at 18-months, and 738/3,827 (19.3%) at 24-months. For much of the pandemic, participants were worried about others not adhering to guidelines, leading to feelings of vulnerability and lack of control.

> *“Not being able to avoid it despite taking sensible precautions. My continuing health effectively depends on other people doing the right things, and that isn’t always going to happen.”*
>
> (P29, female, age 41-50, pre-existing condition(s), baseline)

Non-compliance was sometimes framed in moral terms, reflecting frustration with perceived irresponsible behaviour.

> *“Spread of the virus by careless, arrogant people who will not follow guidelines.”*
>
> (P30, male, age 71-80, no pre-existing condition, 12 months)

Early concerns focused on the lack of testing, vaccines, and treatments, which heightened perceived risk for vulnerable individuals.

> *“I’m worried for my parents and vulnerable friends who may not be able to pull through if they catch it as there is no vaccine or therapeutic drugs available at the moment”*
>
> (P31, male, age 31-40, no pre-existing conditions, baseline)

> *“Lack of testing - nobody knows the real number of cases. I work with a lot of elderly and vulnerable patients and can’t get tested. I’m currently self-isolating due to high temp but will never know if I contracted Covid 19.”*
>
> (P32, female, age 18-30, no pre-existing conditions, baseline)

Later in the pandemic, concerns shifted towards the removal of free testing and broader surveillance systems, alongside worries about new variants and vaccine effectiveness.

> *“I am concerned about the UK government’s action in ending free testing and many of the data collections and studies that may leave us vulnerable to new, more harmful variants. Vaccine inequality across the world is both unjust and dangerous for us all.”*
>
> (P35, female, age 61-70, no pre-existing condition, 24 months)

> *“Also, as time passes the increased likelihood of new variants reducing the effectiveness of vaccines and resulting in more hospitalizations and deaths.”*
>
> (P36, male, age 61-70, no pre-existing conditions, 12 months)

Workplace exposure was another recurring theme, initially among those in high-risk roles and later among those required to return to shared work environments.

> *“I work in a school with children aged 13 and over who have learning disabilities, don’t know how to distance, need personal care, without being allowed to wear a face covering, all the precautions that are insisted upon in other workplaces. How can I protect myself and others.”*
>
> (P37, female, age 41-50, pre-existing condition, 3 months)

> *“Being forced to return to the office in a hot desk environment with colleagues whose children keep bringing Covid home from school.”*
>
> (P38, male, age 18-30, pre-existing condition(s), 24 months)

Policy changes reducing protections were particularly concerning at major transition points, especially for clinically vulnerable individuals.

> *“As a person who is shielding, it is worrying if shielding is lifted, I do not think it is safe to go out into shops etc. Also seeing all the crowds where people are demonstrating, going to the beach and drinking is very worrying, it almost feels like a slap across the face towards people who are following the rules and guidelines.”*
>
> (P39, female, age 41-50, pre-existing condition, three months)

> *“Current increasing rates of infection, hospitalisation and death - at a time when restrictions are being more or less entirely dropped. Clinically vulnerable and extremely vulnerable people are given very little extra help to keep themselves free of the virus now.”*
>
> (P40, female, age 51-60, pre-existing condition, 24 months)

A minority also expressed concerns about the negative societal effects of increased restrictions, particularly for children and the economy.

> *“I am only worried that [my children’s] education might be further disrupted if there is another wave and lockdown occurs again.”*
>
> (P41, female, age 51-60, no pre-existing condition, three months)

> *“I don’t think blanket lockdowns work, just destroying the economy. If you need to shield, then do so, it’s been a knee jerk hysterical reaction.”*
>
> (P42, male, age 41-50, pre-existing conditions, 12 months)

### Government and media

Concerns about government and media centred on trust, reliability of information, and proportionality of risk communication. At baseline, 360/11,113 (3.2%) respondents had one or more concerns about government and/or media, 235/7,048 (3.3%) at 3-months, 237/5,535 (4.2%) at 12-months, 122/4,242 (2.9%) at 18-months, and 94/3,827 (2.5%) at 24-months.

Some participants viewed the government response as inadequate or poorly prepared, contributing to preventable harm.

> *“That our NHS is so ill equipped to face this crisis is unforgivable. These people are the best of all of us and governments have repeatedly treated them shabbily.”*
>
> (P43, female, age 71-80, pre-existing condition(s), baseline)

> *“That it’s [COVID-19] still rife and causing terrible health and social problems and the UK government are basically just ignoring it.”*
>
> (P44, female, age 41-50, pre-existing condition(s), 24 months)

Others felt that government measures were excessive and economically damaging.

> *“Their hysteria and public opinion turning against the government was why a very sensible plan was abandoned in favour of useless draconian measures, which will be pointless as when we come out of isolation everyone will catch the bug again, we might as well get it over with rather than destroy the economy, people’s careers, lose houses etc through these stupid economy harming measures, which ultimately will lead to far more deaths as with the economy depressed the NHS and social care will get less money and people will die, in far greater numbers than the COVID”*
>
> (P45, female, age 31-40, no pre-existing conditions, baseline)

Discrepancies between official messaging and personal experiences generated anxiety and confusion.

> “The government messaging makes me feel really anxious and paranoid, but at the same time those adverts are at odds with my own experience and the experience of most of the people I know who’ve had it (some moderately ill). I know a lot of people who’ve had it now, but no-one has had it to the level advertised on the official government adverts. The difference between my real-life experience and the news really messes my head up.”
>
> (P46, female, age 51-60, no pre-existing condition, 12 months)

Trust was further undermined by perceived rule-breaking among high-profile figures and inconsistencies between UK nations, which were seen to reduce public compliance and create injustice.

> *“Once there was the debacle surrounding Dominic Cummings and his arrogance regarding the regulations and spin he put on them, until then the local community had been excellent at following the guidelines (which were not hard to understand) following that and the total disregard shown by Boris to the rest of us people started going out more not socially distancing as they should. If challenged, they would just say they were ‘doing a Dominic’. Because of this a lot of friends and family do not feel as safe regarding lockdown. The feelings of everyone I know is that there is one rule for them and the rest of us don’t matter”*
>
> (P47, female, age 61-70, no pre-existing condition, 3 months)

> *“The lack of cohesion between the four nations of the UK in managing the response. This has led to significant confusion which in turn has contributed to the success of the virus in spreading. The total disregard by large numbers of people who gathered against the guidance at the time and the softly, softly approach to them by the authorities. They knew what they were doing was wrong, but they also knew there would be no comeback on them.”*
>
> (P48, male, age 61-70, pre-existing condition(s), 18 months)

Media-related concerns often focused on excessive, contradictory, or alarmist coverage that heightened anxiety.

> *“Media scaremongering and contradictory information that confuses, scares and is generally unhelpful in managing the virus.”*
>
> (P48, female, age 41-50, no pre-existing condition, 3 months)

> *“It is impossible to get away from it. Even if you try. The TV will remind you of your duty.”*
>
> (P49, female, age 71-80, pre-existing condition(s), 3 months)

Participants frequently viewed government and media concerns as interconnected, with media perceived to amplify political messaging and policy differences.

> *“Too many politicians do not get the impact of the virus, they put their personal gain in terms of media presence over the health of the nation. Calling for pubs to open may get a few headlines but the deaths and illness it could cause would hit families and the economy hard.”*
>
> (P50, male, age 61-70, no pre-existing condition, 12 months)

> *“My main concern is the lack of clear information especially when England and Wales differ so much, but we hardly ever hear about Wales and their plans in the media.”*
>
> (P51, female, age 71-80, no pre-existing condition, 3 months)

### General concerns

General concerns fell into three areas: timeline, uncertainty, and non-specific worries. These types of concern decreased as the pandemic progresses, with 641/11,113 (5.8%) respondents at baseline reporting a concern in this category, 278/7,048 (3.9%) at 3-months, 218/5,535 (3.9%) at 12-months, 204/4,242 (4.8%) at 18-months, and 82/3,827 (2.1%) at 24-months. Timeline concerns focused on how long the pandemic would last and whether life would return to normal.

> *“Will it ever go away or is it something we have to live with for the rest of our lives, and how our lives will be affected if a vaccine is not found. How long will it take to find a vaccine. Will we ever be able to live a normal life again.”*
>
> (P52, female, age 61-70, pre-existing condition, 3 months)

Uncertainty about the virus, its impacts, and future developments also generated anxiety, particularly regarding unknown long-term consequences and adherence to guidance.

> *“It is the fear of the unknown and anticipation of what is to come. There is a large elderly community here and I worry about them. I worry about the people not adhering to government guidance who undermine the efforts to curtail the spread. I’m worried we aren’t being told the truth about the virus. I worry that no one really knows enough about this virus to understand it and what its future impact will be.”*
>
> (P53, female, age 41-50, no pre-existing conditions, baseline)

Some responses expressed broader, non-specific worries about disrupted life and general health concerns that did not fit other categories, for example “*How it has ruined normal life*” (P54, female, age 71-80, pre-existing condition(s), 3 months), or “*general health concerns*” (P55, female, age 61-70, no pre-existing condition, 18 months).

## Discussion

The findings of this study highlight that concerns about COVID-19 were complex and dynamic, evolving over the course of the pandemic as people attempted to balance the need to protect themselves and others from infection, with concerns about the profound impact of the government pandemic response on daily life and society and the economy more broadly. Overall, the proportion of people reporting concerns decreased over the course of the pandemic, with concerns about personal harm being common throughout. The specific concerns reported changed over the course of the pandemic, shifting in emphasis from pro-social concerns about harm to others during the early stages to more individual-focused worries about personal harm and concerns about others not following rules during the later stages. Concerns about the social and economic impact of the pandemic also decreased over time. These findings indicate a need for ongoing holistic assessment of concerns during pandemics that extend beyond perceptions of personal harm. To fully understand how competing needs and impulses are balanced to determine infection prevention behaviour, conceptualisation and measurement of affective risk appraisal also need to incorporate concerns about harm to others, social and economic impacts, appropriateness and adherence with measures to control the spread of infection, and trust in government and media. This more nuanced understanding of concerns is essential in enabling adaptive, tailored and targeted public health interventions and communication strategies to be rapidly developed and implemented in future pandemics.

The findings of this study indicated that COVID-19 risk perception was underpinned by a wide range of concerns, which is consistent with previous research (39). During the pandemic, governments largely relied on policies relating to social isolation and distancing - particularly before vaccinations were widely available – but these interventions have a considerable psychological and socio-economic cost and disproportionally affect disadvantaged populations (40). PRIME Theory would predict that motivation to engage in a behaviour involves balancing needs and impulses at a particular moment in time (12), and our findings support this view. Concerns about protection for others in people’s immediate social networks and more broadly were prominent in our study during the early stages of the pandemic. This may underpin the association between COVID-19 risk perception and engagement in pro-social behaviour that has been consistently found across several countries (4).

The risk of serious harm had reduced in the UK by the later stages of this study due to a combination of population immunity, widespread vaccination, and differences in the dominant virus variant, but community cases, hospitalisations and deaths still posed a significant risk to population health (41, 42). This corresponded with an overall reduction in the proportion of people in our study reporting concerns by the 24-month follow-up point, but many people remained worried. Concerns often centred about personal harm from COVID-19, compounded by perceived clinical vulnerability, worries about the impact of removal of protection measures, and a perceived decline in infection-prevention behaviour amongst members of the public. This left many of those who remained at risk from harm feeling vulnerable and forgotten by government and society more broadly. The overall decline in the number of people reporting concerns about COVID-19, despite the ongoing harm to public health, may have been indicative of a pandemic fatigue effect, reflecting demotivation to follow protective behaviours that emerges gradually over time, affected by a range of emotions, experiences and perceptions (43). However, this may also have been influenced by a marked change in government policy at this time, accompanied by a shift in messaging from the need for restrictions and personal responsibility to ‘Living with COVID-19’ alongside other respiratory infections (44).

Previous research has highlighted the impact that low trust in government, social trust and individualistic worldviews can have on the acceptance of pandemic mitigation strategies (45, 46). However, these findings are not universal; high trust in government can potentially decrease engagement in preventative behaviour due to an increased sense of security (39, 47). Our study indicated that lack of trust in government, particularly following high-profile examples of government and associated parties breaking lockdown rules without repercussion, was particularly problematic. This impacted on people’s sense of social justice and willingness to engage with policies that were having a major impact on their civil liberties and daily lives. Perceptions of others not following rules and guidelines and lack of enforcement undermined people’s motivation and reduced their sense of control over preventing infection. Conversely, some people felt that government restrictions disproportionate given their perceived low risk of COVID-19 disease. A review of the UK government’s public health response concluded that public health messaging and government communication was often incoherent and inconsistent, particularly in England (48). The findings of our study are consistent with this, reinforcing the need to build trust, ensure that authority figures and institutions role model appropriate behaviour, and to provide coherent and consistent messaging (49).

Exposure to media can influence motivation to engage in infection-prevention behaviour and mental health outcomes, depending on perceived accuracy of the information, trust in sources of information, and type of perceived threat (health risk vs. political risk)(7, 50–52). In this study, people’s views of the government and media were intertwined, with concerns centred around a perceived discrepancy between information presented in mainstream media and people’s lived experiences, as well as perceptions that the quantity of coverage was excessive and provoked anxiety. Social media did not feature prominently in the specific concerns reported in this study, which is consistent with the quantitative findings of the COPE study which indicated that participants considered mainstream media to be more reliable than social media throughout the pandemic (6). Our findings reinforce the importance of ensuring that communication strategies focusing on mainstream media coverage are well planned, consistent and accurate so that members of the public can have confidence in the information that they receive through these channels.

### Strengths and limitations

This study provided novel insights into the evolution of specific concerns that underpinned COVID-19 affective risk perception. A rigorous and in-depth analysis of over 40,000 free-text comments from a large-scale longitudinal prospective study were included in the analysis. Limitations of the study were that the COPE cohort had a higher proportion of older adults and people with long-term conditions relative to the Welsh and UK general population, and people from ethnic minority communities were under-represented. As such, participants were self-selecting and while the data enables us to characterise the experiences of this cohort in detail, and research during future pandemics needs to ensure that the views of under-served groups are fully investigated. Observer bias was minimised surveys were self-completed without a researcher present. The free-text responses asked people about their main concerns rather than all concerns, and as such worries in some categories may have been under-reported.

## Conclusions

This study highlighted the dynamic and multi-dimensional nature of concerns during the COVID-19 pandemic. Our findings reinforce the need for longitudinal research during pandemics to allow changes in concerns to be tracked over time to provide insight into population-level adaptation and inequalities in response to pandemics, highlighting where there may be a need for tailored interventions for specific groups (46, 53, 54). This indicates the need for holistic assessments of concerns that enable competing needs, impulses and fears to be considered in designing equitable and acceptable public health measures.

## Data Availability

Individual-level data from our COPE online survey and qualitative data will not be made publicly available due to data security and ethical considerations. The data provided are of a detailed and sensitive nature. Our public contributors expressed concerns about privacy and security during the development and recruitment stages of this research and did not feel that it was appropriate for individual-level data to be made publicly available. Anonymized data from the COPE study can be made available by the authors on reasonable request, subject to approval from the COPE Study Management Group and Cardiff Metropolitan University Applied Psychology Ethics Panel.

## Supporting information

S1 Table: Demographic characteristics of the COPE cohort at each survey time point

S2 Table: Example quotes for sub-categories at each survey time point

S3 Table: STROBE checklist

## Acknowledgements

We are grateful to our patient and public involvement members for their invaluable contributions to designing and steering this research. This study was facilitated by HealthWise Wales, a Health and Care Research Wales initiative led by Cardiff University in collaboration with SAIL, Swansea University. We are grateful for their invaluable support and expertise. We would like to thank Cardiff Metropolitan University, Cardiff University, PRIME Centre Wales, and Swansea University who have all been immensely supportive of this work, allowing our team the time, resources and infrastructure to get the study up and running quickly during the very early stages of the pandemic.

## References

1. Bish A, Michie S. Demographic and attitudinal determinants of protective behaviours during a pandemic: A review. British Journal of Health Psychology. 2010;15(4):797–824.

2. Wise T, Zbozinek TD, Michelini G, Hagan CC, Mobbs D. Changes in risk perception and self-reported protective behaviour during the first week of the COVID-19 pandemic in the United States. R Soc Open Sci. 2020;7(9):200742.

3. Yang S, Cho S-I. Middle East respiratory syndrome risk perception among students at a university in South Korea, 2015. American Journal of Infection Control. 2017;45(6):e53–e60.

4. Dryhurst S, Schneider CR, Kerr J, Freeman ALJ, Recchia G, van der Bles AM, et al. Risk perceptions of COVID-19 around the world. Journal of Risk Research. 2020;23(7-8):994–1006.

5. Phillips R, Gillespie D, Hallingberg B, Evans J, Taiyari K, Torrens-Burton A, et al. Perceived threat of COVID-19, attitudes towards vaccination, and vaccine hesitancy: A prospective longitudinal study in the UK. Br J Health Psychol. 2022;27(4):1354–81.

6. Phillips R, Hallingberg B, Torrens-Burton A, Wood F, Gillespie D, Elmi-Glennan C, et al. Are you afraid of COVID-19? Motivation and engagement in infection-prevention behaviour in a UK community cohort during the first two-years of the COVID-19 pandemic. . British Journal of Health Psychology. 2025;30(e70034):1–18.

7. Schneider CR, Dryhurst S, Kerr J, Freeman ALJ, Recchia G, Spiegelhalter D, et al. COVID-19 risk perception: a longitudinal analysis of its predictors and associations with health protective behaviours in the United Kingdom. Journal of Risk Research. 2021;24(3-4):294–313.

8. Gibson Miller J, Hartman TK, Levita L, Martinez AP, Mason L, McBride O, et al. Capability, opportunity, and motivation to enact hygienic practices in the early stages of the COVID-19 outbreak in the United Kingdom. Br J Health Psychol. 2020;25(4):856–64.

9. Venkatesan S, Nguyen-Van-Tam JS, Siebers PO. A novel framework for evaluating the impact of individual decision-making on public health outcomes and its potential application to study antiviral treatment collection during an influenza pandemic. PLoS One. 2019;14(10):e0223946.

10. Nibali L, Ide M, Ng D, Buontempo Z, Clayton Y, Asimakopoulou K. The perceived impact of Covid-19 on periodontal practice in the United Kingdom: A questionnaire study. J Dent. 2020;102:103481.

11. Spence JC, Rhodes RE, McCurdy A, Mangan A, Hopkins D, Mummery WK. Determinants of physical activity among adults in the United Kingdom during the COVID-19 pandemic: The DUK-COVID study. Br J Health Psychol. 2020.

12. West R, Michie S, Rubin GJ, Amlôt R. Applying principles of behaviour change to reduce SARS-CoV-2 transmission. Nat Hum Behav. 2020;4(5):451–9.

13. Michie S, van Stralen MM, West R. The Behaviour Change Wheel: a new method for characterising and designing behaviour change interventions. Implementation Science. 2011;6(42).

14. Slovic P, Finucane ML, Peters E, MacGregor DG. Risk as analysis and risk as feelings: Some thoughts about affect, reason, risk and rationality. The feeling of risk: Routledge; 2013. p. 21–36.

15. Tagini S, Brugnera A, Ferrucci R, Mazzocco K, Compare A, Silani V, et al. It won’t happen to me! Psychosocial factors influencing risk perception for respiratory infectious diseases: A scoping review. Applied Psychology: Health and Well-Being. 2021;13(4):835–52.

16. West R, Brown J. Theory of addiction. 2nd Edition ed. Chichester: Addiction Press; 2013.

17. Loewenstein GF, Weber EU, Hsee CK, Welch N. Risk as feelings. Psychol Bull. 2001;127(2):267–86.

18. Savadori L, Lauriola M. Risk perceptions and COVID-19 protective behaviors: A two-wave longitudinal study of epidemic and post-epidemic periods. Social Science & Medicine. 2022;301:114949.

19. Dyer ML, Sallis HM, Khouja JN, Dryhurst S, Munafò MR. Associations between COVID-19 risk perceptions and mental health, wellbeing, and risk behaviours. Journal of Risk Research. 2022;25(11-12):1372–94.

20. Alqahtani MMJ, Arnout BA, Fadhel FH, Sufyan NSS. Risk perceptions of COVID-19 and its impact on precautionary behavior: A qualitative study. Patient Educ Couns. 2021;104(8):1860–7.

21. Cipolletta S, Andreghetti GR, Mioni G. Risk Perception towards COVID-19: A Systematic Review and Qualitative Synthesis. International Journal of Environmental Research and Public Health. 2022;19(8):4649.

22. Kollmann J, Kocken PL, Syurina EV, Hilverda F. The role of risk perception and affective response in the COVID-19 preventive behaviours of young adults: a mixed methods study of university students in the Netherlands. BMJ Open. 2022;12(1):e056288.

23. Leung DYL, Hwu H, Khan S, Mamuji A, Rozdilsky J, Chu T, et al. Understanding the Risk of Social Vulnerability for the Chinese Diaspora during the COVID-19 Pandemic: A Model Driving Risk Perception and Threat Appraisal of Risk Communication-A Qualitative Study. Int J Environ Res Public Health. 2024;21(4).

24. Siddiqui S, Qamar AH. Risk Perception and Protective Behavior in the Context of COVID-19: a Qualitative Exploration. Asian Bioeth Rev. 2021;13(4):401–20.

25. World Health Organisation. Risk communication and community engagement readiness and response to coronavirus disease (COVID-19): interim guidance, 19 March 2020. Geneva; 2020.

26. Bavel JJV, Baicker K, Boggio PS, Capraro V, Cichocka A, Cikara M, et al. Using social and behavioural science to support COVID-19 pandemic response. Nat Hum Behav. 2020;4(5):460–71.

27. World Health Organization Regional Office for Europe. Survey tool and guidance: rapid, simple, flexible behavioural insights on COVID-19: 29 July 2020. Copenhagen; 2020 2020. Contract No.: WHO/EURO:2020-696-40431-54222.

28. Fancourt D, Steptoe A, Wright L. The Cummings effect: politics, trust, and behaviours during the COVID-19 pandemic. Lancet. 2020;396(10249):464–5.

29. Pierce M, Hope H, Ford T, Hatch S, Hotopf M, John A, et al. Mental health before and during the COVID-19 pandemic: a longitudinal probability sample survey of the UK population. Lancet Psychiatry. 2020;7(10):883–92.

30. Hsieh H-F, Shannon SE. Three Approaches to Qualitative Content Analysis. Qualitative Health Research. 2005;15(9):1277–88.

31. Wang Z, Luo S, Xu J, Wang Y, Yun H, Zhao Z, et al. Well-being reduces COVID-19 anxiety: A three-wave longitudinal study in China. Journal of Happiness Studies. 2021;22:3593–610.

32. Mei Y, Tan L, Yang W, Luo J, Xu L, Lei Y, et al. Risk perception and gratitude mediate the negative relationship between COVID-19 management satisfaction and public anxiety. Scientific Reports. 2023;13(1):3335.

33. Wambua J, Loedy N, Jarvis CI, Wong KLM, Faes C, Grah R, et al. The influence of COVID-19 risk perception and vaccination status on the number of social contacts across Europe: insights from the CoMix study. BMC Public Health. 2023;23(1):1350.

34. Hallingberg B, Williams D, Cannings-John R, Hughes K, Torrens-Burton A, Gillespie D, et al. Protocol for a longitudinal mixed-methods study of psychosocial determinants of health behaviour, health and well-being outcomes during the COVID-19 pandemic: The UK COVID-19 Public Experiences (COPE) Study. Figshare: Cardiff Metropolitan University; 2021.

35. Phillips R, Taiyari K, Torrens-Burton A, Cannings-John R, Williams D, Peddle S, et al. Cohort profile: The UK COVID-19 Public Experiences (COPE) prospective longitudinal mixed-methods study of health and well-being during the SARS CoV2 coronavirus pandemic. PLoS One. 2021;16(10):e0258484.

36. Fereday J, Muir-Cochrane E. Demonstrating Rigor Using Thematic Analysis: A Hybrid Approach of Inductive and Deductive Coding and Theme Development. International Journal of Qualitative Methods. 2006;5(1):80–92.

37. Hurt L, Ashfield-Watt P, Townson J, Heslop L, Copeland L, Atkinson MD, et al. Cohort profile: HealthWise Wales. A research register and population health data platform with linkage to National Health Service data sets in Wales. BMJ Open. 2019;9(12):e031705.

38. Giesen L, Roeser A. Structuring a Team-Based Approach to Coding Qualitative Data. International Journal of Qualitative Methods. 2020;19:1609406920968700.

39. Evensen D, Warren G, Bouder F. Satisfaction With Governmental Risk Communication Both Increases and Decreases COVID-19 Mitigation Behaviours. Int J Public Health. 2023;68:1604966.

40. Brooks SK, Webster RK, Smith LE, Woodland L, Wessely S, Greenberg N, et al. The psychological impact of quarantine and how to reduce it: rapid review of the evidence. Lancet. 2020;395(10227):912–20.

41. Office for National Statistics. How coronavirus (COVID-19) compares with flu and pneumonia as a cause of death. 2022.

42. UKHSA data dashboard: COVID-19 Archive data download [Internet]. 2024. Available from: https://ukhsa-dashboard.data.gov.uk/covid-19-archive-data-download.

43. World Health Organisation. Pandemic fatigue – reinvigorating the public to prevent COVID-19: policy framework for supporting pandemic prevention and management. Regional Office for Europe.; 2020.

44. UK Government. COVID-19 response: Living with COVID-19 2022 [Available from: https://www.gov.uk/government/publications/covid-19-response-living-with-covid-19.

45. Siegrist M, Bearth A. Worldviews, trust, and risk perceptions shape public acceptance of COVID-19 public health measures. Proceedings of the National Academy of Sciences. 2021;118(24):e2100411118.

46. Hanna K, Clarke P, Woolfall K, Hassan S, Abba K, Hajj TE, et al. The perception of risk in contracting and spreading COVID-19 amongst individuals, households and vulnerable groups in England: a longitudinal qualitative study. BMC Public Health. 2023;23(1):653.

47. Liu S, Zhu J, Liu Y, Wilbanks D, Jackson JC, Mu Y. Perception of strong social norms during the COVID-19 pandemic is linked to positive psychological outcomes. BMC Public Health. 2022;22(1):1403.

48. British Medical Association. BMA Covid Review 4: The public health response by UK governments to COVID-19. 2022.

49. Warren GW, Lofstedt R. Risk communication and COVID-19 in Europe: lessons for future public health crises. Journal of Risk Research. 2022;25(10):1161–75.

50. Oh SH, Lee SY, Han C. The Effects of Social Media Use on Preventive Behaviors during Infectious Disease Outbreaks: The Mediating Role of Self-relevant Emotions and Public Risk Perception. Health Commun. 2021;36(8):972–81.

51. Erfei Z, Qiao W, Eileen MC, Jennifer AA. Media trust and infection mitigating behaviours during the COVID-19 pandemic in the USA. BMJ Global Health. 2020;5(10):e003323.

52. Allington D, Duffy B, Wessely S, Dhavan N, Rubin J. Health-protective behaviour, social media usage and conspiracy belief during the COVID-19 public health emergency. Psychological Medicine. 2021;51(10):1763–9.

53. Stechemesser A, Kotz M, Auffhammer M, Wenz L. Prolonged exposure weakens risk perception and behavioral mobility response: Empirical evidence from Covid-19. Transportation Research Interdisciplinary Perspectives. 2023;22:100906.

54. Reme B-A, Wörn J, Skirbekk V. Longitudinal evidence on the development of socioeconomic inequalities in mental health due to the COVID-19 pandemic in Norway. Scientific Reports. 2022;12(1):3837.

